# The efficacy of omalizumab treatment in patients with non-atopic severe asthma

**DOI:** 10.1101/2022.01.29.22270075

**Authors:** Emel Atayik, Gökhan Aytekіn

**Author notes:** **Correspending authors Address for reprints: Emel Atayik**, Allergy and Clinical İmmunology Specialist, University of Health Sciences Konya City Hospital, Division of Allergy and Clinical Immunology, Konya / TURKEY. **Informed consent** The study protocol was approved by the Ethics Committee of Necmettin Erbakan University, Meram Faculty of Medicine (meeting date: January 21, 2022 and decision no: 2022/3608). **Funding source:** No funding was secured for the study. **Financial disclosure:** The authors have no financial relationship relevant to this article to disclose. **Disclaimers:** None. Manuscripts that have not been presented orally or as a poster.

## Abstract

**Introduction:** Non-atopic asthmatic patients generally have more severe clinical manifestations frequently accompanied by chronic sinusitis and nasal polyps, with more limited therapeutic options as compared to atopic asthmatic patients. This has led to a search for novel therapeutic approaches including omalizumab for non-atopic asthmatic patients who are not adequately controlled with current therapies.

**Materials and Methods:** In this retrospective study undertaken between 1st August 2020 and 31st December 2021 at a tertiary allergy clinic, data from 61 non-atopic asthmatic patients inadequately controlled with optimum therapy was examined.

**Results:** A total of 61 patients with severe asthma were included in the study [Female: 48 (78.7%), male: 13 (21.3%], mean age: 49 (18-71) years). The mean duration of asthma was 60 (18-160) months. A statistically significant increase in FEV1 (Forced expiratory volume in one second), FVC (forced vital capacity), and ACT (asthma control test) scores were found after 1 year of omalizumab treatment (p < 0.001, for all parameters). Omalizumab treatment was associated with a significant decrease in the number of asthma exacerbations, asthma-related hospitalization, duration of hospitalization, and the number of unplanned emergency room visits after 1 year (p < 0.001, for all parameters). One year treatment with omalizumab led to significant changes in eosinophil counts and serum IgE levels as well (p < 0.001 and p< 0.001)

**Conclusion:** In the atopic severe asthma patient group omalizumab treatment provided similar clinical benefits to those observed in patients with severe atopic asthma, suggesting that it may be a useful therapeutic option in patients with non-atopic asthma who failed to benefit from step 4 and step 5 treatments.

## Introduction

Omalizumab is a humanized recombinant anti-immunoglobulin (Ig) E monoclonal antibody used for the treatment of several allergic disorders (1-3). It binds to IgE heavy-chain, thereby reducing free IgE levels and preventing it from binding to its receptors on mast cells and basophils. Omalizumab has been approved for use in severe persistent allergic asthma and chronic urticaria resistant to antihistamines. Also, based on the central role of IgE in the pathogenesis of many allergic disorders, “off-label” use of omalizumab has been a common practice, mainly for anaphylaxis, food allergies, and drug allergies, but also for other conditions such as allergic rhinitis, allergic bronchopulmonary aspergillosis, atopic dermatitis, nasal polyps, and Churg-Strauss syndrome (4).

The 2021 Global Initiative for Asthma (GINA) guidelines recommend omalizumab treatment for patients with moderate to severe asthma who are not controlled with step 4 or 5 treatments (5). On the other hand, use of omalizumab in asthmatic patients is limited to atopic asthmatic subjects, and the presence of atopy should be confirmed by demonstrating IgE-mediated sensitivity against at least one perennial allergen using skin prick or in vitro testing (6). However, both atopic and non-atopic asthma patients exhibit a similar immunologic profile, including the elevation of cytokines such as IL-4 and IL-13, increased production of IgE, and increased expression of Fc-epsilon-RI (FceRI) mRNA. Furthermore, a multitude of studies have directly or indirectly shown that IgE plays a role in asthma pathogenesis independent of atopy (7). For instance, a 5-fold increased occurrence of asthma has been detected in non-atopic subjects with elevated total serum IgE levels (8). Non-atopic asthmatic patients generally have more severe clinical manifestations frequently accompanied by chronic sinusitis and nasal polyps, with more limited therapeutic options as compared to atopic asthmatic patients. This has led to a search for novel therapeutic approaches including omalizumab for non-atopic asthmatic patients who are not adequately controlled with current therapies. Studies of omalizumab in non-atopic asthmatic patients are scarce in number and generally include case reports (9-11) or studies with relatively smaller sample size (7, 10, 12). Thus, in this retrospective study we decided to examine the effect of omalizumab on clinical and laboratory parameters in a group of patients with non-atopic severe asthma who received at least 1 year of omalizumab treatment.

## Materials and Methods

In this retrospective study undertaken between 1^st^ August 2020 and 31^st^ December 2021 at our Allergy and Immunology Unit of a Tertiary Center, data from 78 non-atopic asthmatic patients inadequately controlled with optimum therapy was examined, following approval for “off-label” use of this agent from the Turkish Drug and Medical Device Agency of the Turkish Ministry of Health. Seventeen patients were excluded due to several reasons (e.g. short duration of omalizumab treatment precluding assessment of efficacy, treatment at another healthcare facility, or missing data in patient files), leaving 61 non-atopic severe asthma patients for review.

Data on age, gender, smoking status, BMI (body mass index, kg/m2), comorbid conditions (nasal polyps, chronic rhinosinusitis, and chronic urticaria), duration of asthma, and duration of omalizumab treatment were retrieved from patient files. Also recorded were pulmonary function test [FEV1 (Forced expiratory volume in one second), FVC (forced vital capacity), and FEV1/FVC] results and ACT (asthma control test) scores prior to initiation of omalizumab treatment, oral corticosteroid dosage (daily) within the past year, and unplanned emergency room visits, number of asthma exacerbations, and hospitalizations in the past year. Pulmonary and clinical parameters were recorded again following 1 year treatment with omalizumab.

Severe asthma was diagnosed based on GINA asthma guidelines (13). A nSpire ZAN 100 spirometer device was used for spirometry measurements.

Abbott Cell Dyn 3700 (Sheath reagent) and Siemens BN II/ BN ProSpec system (particle-enhanced immunonephelometry) were used for automated complete blood counts and quantitative determination of serum immunoglobulin (Ig) E, respectively.

Patients underwent allergy testing using a skin-prick panel of 24 inhaled aeroallergens from 8 classes (dog, cat, dust mite, grass, tree, ragweed, mold, and cockroach) (ALK, Abello, Madrid, Spain). A wheal diameter of>5 mm with flare at 20 minutes was considered a positive result, and patients were considered as non-atopic, in the absence of any reaction to any allergens or in the absence of allergen-specific IgE (14).

Omalizumab (Xolair; Novartis, Basel, Switzerland) dosage was determined using the standard dozing scheme based on total serum IgE and bodyweight. Asthma was considered well controlled with an ACT score of ≥20, partially controlled with an ACT score of 15-19, and poorly controlled with an ACT score of <15 (15).

The study protocol was approved by the Ethics Committee of Necmettin Erbakan University, Meram Faculty of Medicine (meeting date: January 21, 2022 and decision no: 2022/3608).

All data obtained and recorded in the study forms were analyzed using IBM SPSS 20.0 (Chicago, IL, USA) statistical software program. The normal distribution of discrete and continuous numerical variables was tested with Kolmogorov-Smirnov test. Descriptive statistics for discrete and continuous numerical variables were expressed as mean ± standard deviation (SD) or median (minimum-maximum), while categorical variables were expressed as the number of cases and (%). Categorical variables were assessed with chi-square test, while continuous variables were assessed using t-test or Mann-Whitney U test. Dependent variables with normal distribution were compared using paired-samples T test, while those without normal distribution were compared with Wilcoxon test. Categorical dependent variables with more than 2 nominal values were compared with McNemar-Bowker test. A p value of less than 0.05 was considered to be statistically significant.

## Results

A total of 61 patients with severe asthma were included in the study [Female: 48 (78.7%), male: 13 (21.3%], mean age: 49 (18-71) years). Mean duration of asthma was 60 (18-160) months. Main comorbidities included chronic rhinosinusitis (57.4%) and nasal polyps (24.6%). Clinical, laboratory and demographic parameters of the asthma patients are summarized in Table 1.

**Table 1:** Clinical, demographic and laboratory parameters in asthma patients

| Parameters | Results (n: 61) |
| --- | --- |
| Age, years | 49 (18-71) |
| Gender, female, n (%) | 48 (78.7) |
| BMI, kg/m<sup>2</sup> | 28.42 ± 6.27 |
| Smoking status, n (%) |  |
| • Never smoked | • 59 (96.8) |
| • Ex-smoker (≥1 year) | • 1 (1.6) |
| • Smoker, n (%) | • 1 (1.6) |
| Duration of asthma, years | 60 (18-160) |
| Duration of omalizumab treatment, months | 16 (12-66) |
| Dose of omalizumab |  |
| • 150 mg/ 4 weeks | • 22 (36.1) |
| • 300 mg/ 4 weeks | • 36 (59.0) |
| • 450 mg/ 4 weeks | • 3 (4.9) |
| Main comorbidities, n (%) |  |
| • Chronic rhinosinusitis | • 35 (57.4) |
| • Nasal polyps | • 15 (24.6) |
| • Chronic urticaria | • 7 (11.5) |
| HRCT findings of lungs |  |
| • Normal | • 23 (37.7) |
| • Emphysematous changes | • 9 (14.8) |
| • Bronchiectasis and ground-glass opacities | • 13 (21.3) |
| • Linear atelectasis | • 10 (16.4) |
| • Parenchymal fibrosis | • 4 (6.6) |
| • Mosaic perfusion | • 2 (3.3) |

BMI: body mass index, HRCT: High-resolution computed tomography

As compared to baseline, a statistically significant increase in FEV1, FVC, and ACT scores was found after 1 year of omalizumab treatment (p < 0.001, for all parameters). After 1 year of omalizumab treatment, there was not a significant change in FEV1/FVC (p: 0.095) (Figure 1) (Table 2). Furthermore, omalizumab treatment was associated with a significant decrease in the number of asthma exacerbations, asthma-related hospitalization, duration of hospitalization, and number of unplanned emergency room visits after 1 year (p < 0.001, for all parameters) (Table 2) (Figure 2).

**Table 2:** Clinical, pulmonary and laboratory parameters in patients with severe asthma patients before and 1 year after omalizumab treatment

| Parameters | Before omalizumab treatment | After 1 year of omalizumab treatment | p |
| --- | --- | --- | --- |
| FEV1, % | 60.71 ± 19.73 | 73.67 ± 21.76 | <0.001 |
| FVC, % | 64.36 ± 76.30 | 76.30 ± 19.85 | <0.001 |
| FEV1/FVC | 80 (46-125) | 81.0 (70-107) | 0.095 |
| ACT scores | 13 (11-17) | 21 (8-25) | <0.001 |
| Unplanned visit to ER, year | 4 (1-12) | 0 (0-12) | <0.001 |
| Number of hospitalizations, | 1 (0-8) | 0 (0-8) | <0.001 |
| Oral corticosteroids, mg/day | 8 (0-16) | 0 (0-11) | <0.001 |
| Duration of hospitalization, day | 5 (0-25) | 0 (0-15) | <0.001 |
| Asthma control |  |  | <0.001 |
| • Well controlled asthma | - | 42 (68.9) |  |
| • Partially controlled asthma | 21 (34.4) | 6 (9.8) |  |
| • Poorly controlled asthma | 40 (65.6) | 13 (21.3) |  |
| Eosinophil count, cell/mm<sup>3</sup>, | 190 (6.50-2470) | 150 (0.3-1800) | <0.001 |
| Serum IgE, IU/ml, | 47.70 (7-310) | 117 (11-1070) | <0.001 |

FEV1: Forced expiratory volume in the first second, FVC: forced vital capacity, ACT: asthma control test, ER: Emergency room,

**Figure 1:**
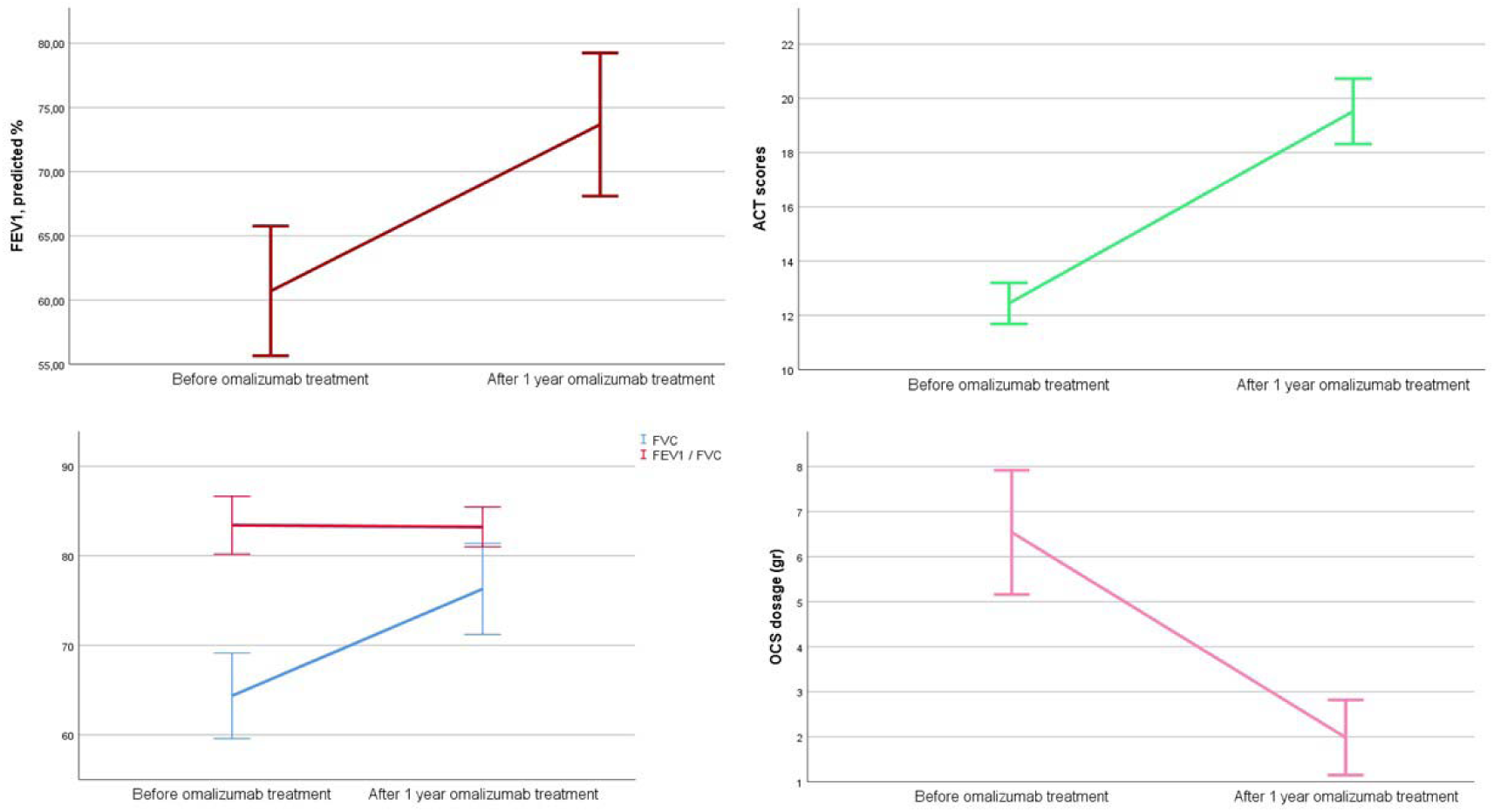
FEV1, FVC, and FEV1/FVC results, ACT scores and daily OCS usage of asthma patients before and 1 year after omalizumab treatment FEV1: Forced expiratory volume in the first second, FVC: forced vital capasity, ACT: asthma control test, OCS: oral corticosteroids

**Figure 2:**
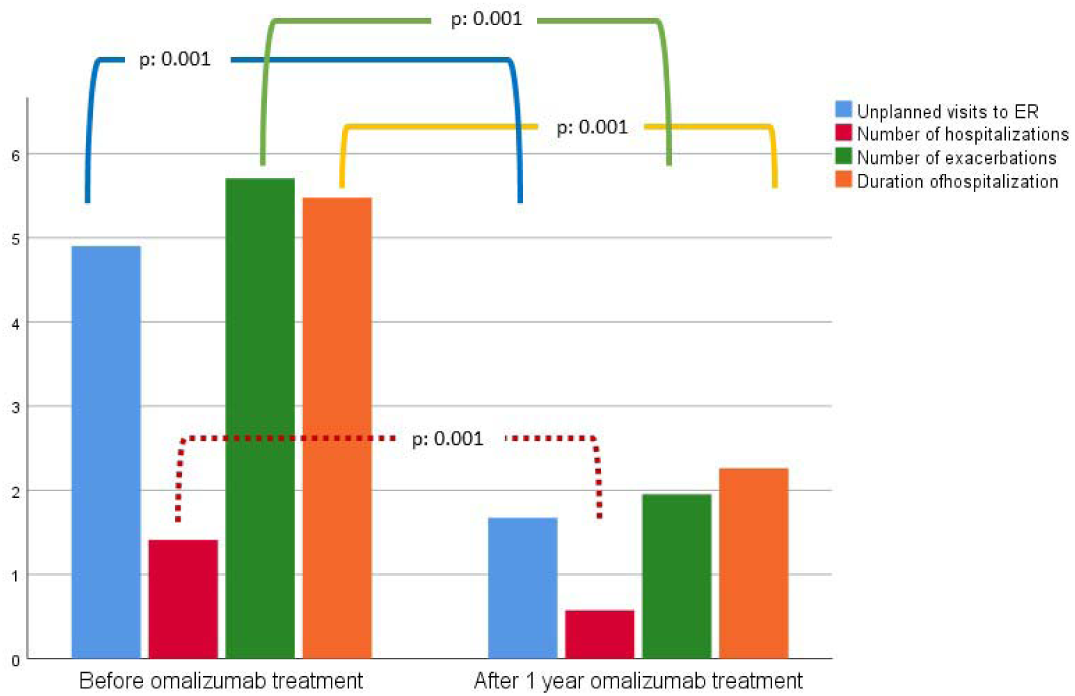
Unplanned visits of ER, number of hospitalizations and exacerbations, and duration of hospitalizations of asthma patients before and 1 year after omalizumab treatment ER: Emergency room

One year treatment with omalizumab led to significant changes in eosinophil counts and serum IgE levels as well (p < 0.001 and p < 0.001) (Figure 3). Table 2 summarizes the changes in clinical and laboratory parameters after 1 year of omalizumab treatment.

**Figure 3:**
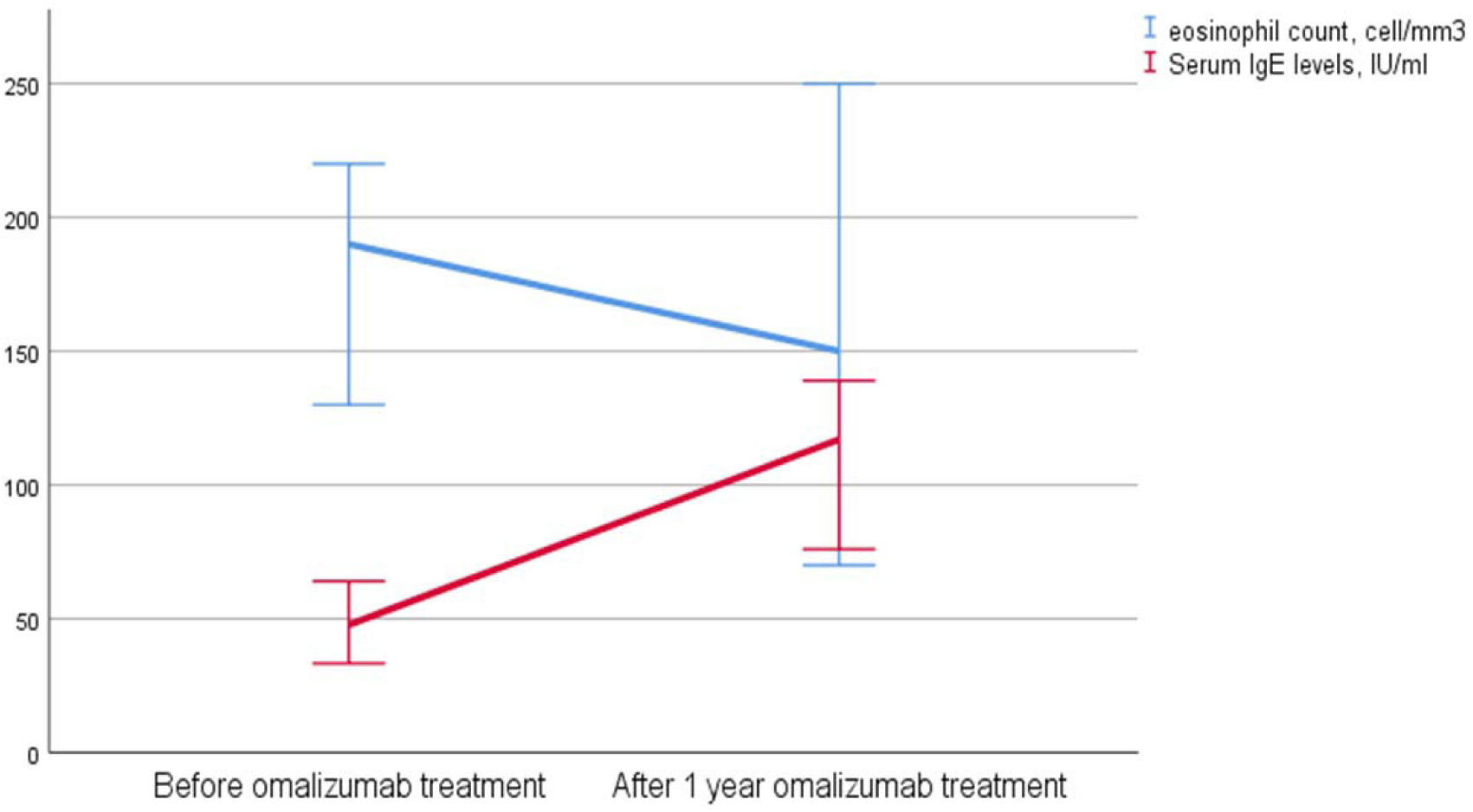
Eosinophil counts and serum IgE levels of asthma patients before and 1 year after omalizumab treatment

## Discussion

Literature data on the efficacy of omalizumab in patients with non-atopic asthma is scarce. In this study evaluating the effect of omalizumab treatment in this patient group, we reached two important conclusions. Firstly, omalizumab was associated with an increase in FEV1, FVC, and ACT scores, and with an decrease in asthma exacerbations, asthma-related hospitalizations, length of hospital stay, and daily need for OCS among 61 patients with non-atopic severe asthma. Secondly, omalizumab treatment led to decrease in eosinophil count and increase in serum total IgE level. These results suggest that omalizumab treatment may offer certain benefits in non-atopic asthmatic patients who are unresponsive to other treatments.

As compared to atopic asthmatic patients, non-atopic asthma patients have fewer therapeutic options, and until now only limited number of studies have assessed omalizumab use in non-atopic asthmatics. In a case report by Menzella et al., a 52-year-old patient with severe non-atopic asthma (receiving formoterol 18 mcg/day; budesonide 640 mcg/day; tiotropium bromide 18 mcg/day and prednisone 25 mg/day), past history of smoking, allergic rhinitis and nasal polyps experienced significant symptomatic improvement with only 16 weeks of treatment with omalizumab (9). Da Llano et al., observed increased FEV1 and ACT scores, and reduced number of asthma exacerbations with omalizumab treatment in 29 non-atopic asthmatic patients, similar to those in atopic patients (7). Again, Sözener et al. reported increase in ACT scores and FEV1 and decrease in asthma exacerbations, asthma-related hospitalizations, and daily oral corticosteroid dose among 13 patients with non-atopic asthma (12). Our results are similar and consistent with those previous observations.

IgE antibodies produced against common aero-allergens represent the main characteristic of atopic asthma. Bridging of IgE molecules on mast cells and basophils by protein allergens results in the activation of these cells, leading to the release of preformed mediators such as histamine, or newly formed mediators such as prostaglandins and leukotrienes from the mast cells. Therefore, both IgE and mast cells play important roles in the pathogenesis of asthma (10). Omalizumab prevents the interaction between IgE and its receptors by forming complexes with free IgE (16). Furthermore, it downregulates the Fc-epsilon-RI receptors found on the cell surface of basophils and mast cells (17). In Berger et al.’s study, incubation of bronchial tissue samples from asthmatic patients with omalizumab resulted in a decrease in specific and non-specific bronchial hyper-responsiveness, which was postulated to be associated with the inhibition of bronchial mast cell degranulation (18).

Although these findings may suffice to explain the efficacy of omalizumab in patients with atopic asthma, other hypotheses have been put forward to account for the efficacy of omalizumab in non-atopic asthma patients. First of these hypotheses involve a reduction in bronchial mucosal IgE positive mast cell numbers (6) and anti-inflammatory effects, leading to clinical benefit (19, 20). DjukanovicR et al. showed a significant reduction in airway inflammation markers assessed with induced sputum and bronchial biopsies following omalizumab treatment (21). Another hypothesis points out to increased release of cytokines such as IL-4 and IL-13 via stimulation of intracellular pathways without cross-linking with allergens, as well as to direct binding of IgE to eosinophils, neutrophils, and monocytes, activating these cells (22, 23). Omalizumab treatment was shown to result in reduced sputum eosinophil counts, as well as a decrease in CD4 + and CD8 + lymphocytes, B lymphocytes, and interleukin-4 positive cells (19, 14); however, a similar mechanism of action is yet to be confirmed in non-atopic asthma patients. Based on this hypothesis, omalizumab is expected to reduce in eosinophil counts by decreasing IL-4 and IL-13 secretion, also in patients with non-atopic asthma. The significant reduction in eosinophil counts after 1 year of omalizumab treatment in our study may be related with this mechanism. Another hypothesis is based on the observation that non-atopic asthmatic patients have more frequent symptoms and more extensive airway obstruction (8), suggesting that all asthma patients have an atopic component and increased IgE may result from a yet-undefined allergen (8, 25). This latter hypothesis should be tested in larger patient samples in order to better elucidate the efficacy of omalizumab in patients with non-atopic asthma.

Our study has certain advantages and limitations. The advantages include a relatively larger sample size and data based on real-life observations. On the other hand, the cross-sectional design represents its major disadvantage. Also, although patients were considered atopic based on skin prick testing and in vitro testing, some patients might have been sensitized through allergens not tested with our study panels. However, since atopy is reduced with age and our patients mostly consisted of middle to advanced age individuals, it may be assumed that some patients with previous allergen sensitivity may have experienced reduced sensitivity over time, leading to their classification in the non-atopic category with subsequent false negativity. Also, since our study only assessed a 1-year period of omalizumab treatment, our results may not be generalized to longer-term efficacy.

In conclusion, in this patient group omalizumab treatment provided similar clinical benefits to those observed in patients with severe atopic asthma, suggesting that it may be a useful therapeutic option in patients with non-atopic asthma who failed to benefit from step 4 and step 5 treatments.

## Data Availability

All data produced in the present study are available upon reasonable request to the authors

## Acknowledgement

None

